# What Information exists about the disease and health systems burdens of lesser studied enteric viral infections in north west Europe? A rapid systematic mapping review

**DOI:** 10.1101/2025.06.09.25329265

**Authors:** Julii Brainard, Xia Wang-Steverding, Isabel Catalina Swindells

## Abstract

**Background:** Availability of multiplex PCR methods means that less studied viruses are often linked to acute gastrointestinal infection. Among the virus genera increasingly linked to illness and health care usage are astroviruses, sapoviruses, enteric adenoviruses and parechoviruses.

**Methods:** A systematic mapping review was undertaken to evaluate availability of published evidence that might be used to describe the disease burden or health care system usage associated with infection status with the target viruses among north west European populations. Availability of information was tabulated with respect to which viruses were tested, specific symptoms, sequelae and indicators of health service usage.

**Results:** At least 49 studies were available that described outcomes related to each of the target viruses. The most commonly documented symptom was presence/absence of diarrhoea. Metrics such as hospitalisation rates were available in a minority of studies, and no data were published about disability-adjusted-life-years, quality-of-life or life expectancy in relation to infection status.

**Conclusions:** It is feasible to estimate typical rates of hospitalisation or length of stay with infection by astroviruses, sapoviruses, enteric adenoviruses or parechoviruses.

## Introduction

In the UK, surveillance is routine for many gastrointestinal pathogens, such as norovirus, E. Coli O157, and Cryptosporidium spp. Establishing the merits of expanding current routine surveillance practices in the UK to include other gastrointestinal pathogens would ideally utilise specific data about the actual or potential disease burden that can be associated with individual infectious agents in the population. With the introduction of multiplex PCR methods for routine diagnosis, several less studied viruses that have been associated with symptomatic gastrointestinal illness are increasingly identified in stool samples in many countries. Among the virus genera that are increasingly documented for their potential links to symptomatic gastrointestinal symptoms as well as potentially severe sequelae are astroviruses [1-3], sapoviruses [4-6], enteric adenoviruses [7-9] or parechoviruses [10-12]. In the remainder of this article, we refer collectively to these four viral genera as ‘target viruses’. The purpose of this rapid mapping review was to document what evidence exists that could be most relevant to UK populations, for possible disease burdens and health care system usage (or not) associated with infections by these target viruses. This information can inform which specific outcomes in existing scientific literature could most feasibly indicate the disease or health care system burden in UK and similar populations caused by the target viruses. The latter information, estimates of disease burden or health care usage, would be useful to inform if surveillance should be increased for infection by the target viruses in the UK or similar settings.

## Methods

To undertake a mapping review, we applied systematic review methods [13] to quickly and relatively comprehensively look for and summarise presence or absence of information about conventional disease burden metrics (such as disability adjusted life years), specific disease outcomes among or health care usage rates by humans who had been tested for infection status with any of the target viruses. Ethics approval was not required to undertake this research because this is a synthesis of data already in the public domain and intended for widespread dissemination.

### Search strategy

Only scientific articles were included, and only data in the existing published articles were extracted. Because this was a rapid mapping review, only published articles that the authors could obtain easily via existing institutional access agreements were included: we did not order articles or request more information from primary study authors.

A bibliography was assembled by JB combining hits from the Pubmed and Scopus search portals. Box 1 lists the searches used for Pubmed. The searches were designed to be certain to find the articles listed in Box S1. The articles in Box S1 were found using Google Scholar searches with parts of the final search phrases and were used to develop the search terms because they seemed likely to contain relevant data. To have the results rely on relatively recent data and the greater reliability of relatively modern diagnostic methods, all included articles had to be published in year 2000 or later and to refer to data collected in the year 1994 or later. Studies published before the year 2000, duplicated studies and animal-only studies were removed by both automatic and manual methods before screening began. Missing abstracts were obtained where publicly available but if an abstract was unavailable then the article was excluded. All articles were screened in Microsoft Excel on title, journal title and abstract independently by XW and ICS, with JB providing a third opinion in case of disagreement at all stages.

**Box 1: Search phrases in respective bibliographic databases**

PUBMED:

((Astrovirus or Sapovirus or adenovirus or Parechovirus) and (infection or case or prevalence or incidence) and (gastrointestinal or gastroenteritis or enterovirus or intestinal or diarrhea or enteric))

SCOPUS: (*also limited to humans only, terms within abstract*.*title*.*keywords*)

((Astrovirus or Sapovirus or adenovirus or Parechovirus) and (infection or case or prevalence or incidence) and (gastrointestinal or gastroenteritis or enterovirus or intestinal or diarrhea or enteric))

**Box S1. Articles used to test search phrases: search phrases had to find these articles**

***Are among final included articles***

Brown *et al*. “Viral gastrointestinal infections and norovirus genotypes in a paediatric UK hospital, 2014– 2015.” *Journal of Clinical Virology* 84 (2016): 1-6.

Jarchow-Macdonald *et al*. “First report of an astrovirus type 5 gastroenteritis outbreak in a residential elderly care home identified by sequencing.” *Journal of Clinical Virology* 73 (2015): 115-119.

***Met many inclusion criteria but outside eligible geographic area***

Amoroso *et al*. “Ten different viral agents infecting and co-infecting children with acute gastroenteritis in Southern Italy: Role of known pathogens and emerging viruses during and after COVID-19 pandemic.” *Journal of Medical Virology* 96.5 (2024): e29679.

Badjo *et al*. “Genetic diversity of enteric viruses responsible of gastroenteritis in urban and rural Burkina Faso.” *PLOS Neglected Tropical Diseases* 18.7 (2024): e0012228.

Bucci *et al*. “Clinical and neurodevelopmental characteristics of Enterovirus and Parechovirus Meningitis in neonates.” *Frontiers in Pediatrics* 10 (2022): 881516.

Chirinda *et al*. “Detection of Enteric Viruses in Children under Five Years of Age before and after Rotavirus Vaccine Introduction in Manhiça District, Southern Mozambique, 2008–2019.” *Viruses* 16.7 (2024): 1159.

Manoha *et al*. “Multisite community-scale monitoring of respiratory and enteric viruses in the effluent of a nursing home and in the inlet of the local wastewater treatment plant.” *Applied and Environmental Microbiology* (2024): e01158-24.

Pitkänen *et al*. “The role of the sapovirus infection increased in gastroenteritis after national immunisation was introduced.” *Acta Paediatrica* 108.7 (2019): 1338-1344.

Salavatiha *et al*. “Investigation the Prevalence of Norovirus, Rotavirus, Human Bocavirus, and Adenovirus in Inpatient Children with Gastroenteritis in Tehran, Iran, During 2021-2022.” *Iranian Journal of Medical Microbiology* 18.4 (2024): 230-237.

Varghese *et al*. “Etiology of diarrheal hospitalizations following rotavirus vaccine implementation and association of enteric pathogens with malnutrition among under-five children in India.” *Gut Pathogens* 16.1 (2024): 22.

Vu *et al*. “Novel human astroviruses: prevalence and association with common enteric viruses in undiagnosed gastroenteritis cases in Spain.” *Viruses* 11.7 (2019): 585.

### Inclusion criteria

Only articles in scientific peer review literature were eligible. Human participants had to have been investigated for possible infection with one of the eligible virus genera: astrovirus, adenovirus, sapovirus or parechovirus. A majority of any identified cases had to be laboratory confirmed. Positivity (or lack thereof) had to be linked to specific symptoms, disease burden metrics or health care usage metrics (or lack thereof). The data had to be used in an original quantitative analysis. Qualitative studies, commentaries, diagnostic or teaching vignettes, theoretical modelling, mechanistic or non-systematic review studies were not eligible. Studies about viral presence or infection effects in animals or environmental samples were ineligible. Primary research data had to be processed on individual participant samples (minimum total persons tested = 20), using exactly one sample per patient rather than multiple samples per patient or unclear sample counts per patient, and the study had to report on at least one of: positivity rates, health care system usage, symptoms or infection sequelae for participants. Tissue samples from recently deceased persons or late gestation foetuses were eligible as indicators of infection/outcome association, as long as the samples could be identified as relevant to exactly one per person.

Although our interest included enteric adenoviruses, defining enteric adenoviruses was not straightforward. Adenoviruses especially associated with respiratory illness have also been linked to gastrointestinal symptoms. Moreover, gastrointestinal infections often cause symptoms outside the gastrointestinal tract; the target viruses in our review are especially associated with meningitis [14, 15] and myocarditis [16, 17] for instance. We did not want to focus our inclusion criteria only on symptoms in the gastrointestinal tract and nor did we want to broaden our review to include disease burden metrics linked to many adenovirus *spp*. that usually cause only respiratory infection symptoms. We therefore opted for a compromise approach, which was to include any infection with one of the target virus genera as long as any non-respiratory symptom(s) were listed among persons who were tested. “Respiratory” for our purposes meant symptoms in the throat, lungs, nasal cavities or ears. We applied this criterion at the earliest review stages (screening abstracts) which means that a study which described in the abstract only respiratory symptoms, was excluded.

Preliminary literature searches indicated that a huge (> 2000) number of studies exist in scientific literature that could indicate something about disease impacts or prevalence of relevant infection or linked health care system usage, linked to infection status with our target viruses. In order to make this mapping review exercise both rapid and feasible with available resources, we restricted study inclusion to populations in countries in north west Europe, specifically countries that border the North Sea or border the UK. These populations had high probability of being similar to UK populations. The specific inclusion criterion was that participants had to be present at time of testing in UK, Ireland, mainland France, Belgium, Netherlands, Germany, Denmark, Norway or Sweden. This geographic grouping was also reasonable because the eligible countries are all high income and broadly have common dietary and lifestyle customs and age and genetic demographics. Most of these eligible countries are or were members of the European Union which means that there have been high rates of population movement between these nations, due to historical factors as well as modern political and economic ties. It is possible that climatic factors also have association with prevalence or seasonality of infection by at least some of the eligible viruses. There is a high degree of climatic similarity within this region, too [18].

## Data extraction and Summary

Box S2 lists the information that was extracted from each included article. A single extractor (either XW or ICS) extracted data for each eligible article. Initially extracted data were confirmed or revised if indicated, by another single extractor (XW or ICS) with JB providing a final decision in case of disagreements.

For astrovirus, sapoviruses or parechoviruses, we did/do not differentiate by serotype within the genus. Risk of bias was not assessed for included studies. Our results are reported as frequency counts and percentages.

**Box S2. Information extracted from each included article**

Bibliographic reference

Year of publication

Study design

Month(s)+Year(s) of data collection

Location (country) of cases/humans investigated

Maximum count of participants in study (count at start of any flow chart/patient selection process diagram)

Were cases (*3 possible answers*) only laboratory-confirmed; mix of laboratory confirmed and clinically defined; unclear

*Questions with yes/no/unclear answers:*

Were majority of participants under age 5 years?

Were majority of participants age 65 years and older?

Were majority of participants in a vulnerable group, describe it (eg HIV+, organ transplant recipients, etc)?

Were most participants clearly tested for (*yes/no answers only possible)*: astrovirus, sapovirus, parechovirus

Were most patients clearly tested for adenovirus? (*possible answers were Not tested; serotypes unclear/mixed; serotype 40/41; serotypes 1-5, 7, 14 or 21; serotypes unclear)*

Were any of these disease experiences, symptoms, disease burden or health care metrics clearly documented for most patients (patient = suspected case)? (*yes/no only possible answers; since most are rates, the denominators were required for yes answers*)

Whether tests were administered (*for each of the eligible viruses; yes/no answer)*

Diarrhea; meningitis; fever; myocarditis (*yes/no answer each)*

Underlying conditions or co-existing vulnerability (*yes/no answer)*

DALY or QALY *(which or both or neither as answers)*

Case fatality rate or complete information needed to calculate CFR (*yes/no answer)*

(healthy) life expectancy (*yes/no answer)*

Presentation rate *(among all persons positive in cohort including in community, yes/no answer)*

Hospitalisation rate *(among presenters, yes/no answer*)

Hospital Bed days (*among confirmed cases, yes/no answer*)

Were treatment details described for any patients? (*yes/no/unclear as possible answers*)

Was information clearly available about what percentage of cases were asymptomatic (*need total case count, yes/no answer*)?

## Results

The count of included articles was 161, describing 162 distinctive study populations (“studies”). Almost half (79, 48.8%) were published after 2012. Tables 1 and 2 provides descriptive statistics about the extracted data in the included studies. Figure 1 shows the study selection process. Supplemental Table 1 (available from authors) contains all included study bibliographic references and the extracted data from each included article.

**Table 1.** Summary of contextual information extracted from included studies.

|  |  |  |  |
| --- | --- | --- | --- |
| ***Years of publication*** |  |  |  |
| 2000-2006 40 (25%) | 2007-2013 43 (27%) | 2014-2019 44 (27%) | 2020-2024 35 (22%) |
| ***Maximum size of patient cohort tested (start of patient recruitment as reported by authors)*** |  |  |  |
| 20-99 persons, 23 (14%) | 100-249 persons, 43 (27%) | 250-999 persons, 49 (30%) | 1000+ persons, 44 (27%) |
| ***Study designs*** |  |  |  |
| Prospective observational<br>91 (56%) | Retrospective<br>observational 66 (41%) | Prospective intervention<br>2 (1.2%) | Other or unclear<br>3 (1.9%) |
| ***Most common countries*** |  |  |  |
| Germany 37 (23%) | Netherlands 35 (22%) | UK 33 (20%) | 58 (36%) only other<br>countries |
| ***Counts of studies where...*** |  |  |  |
| Majority of participants<br>were age < 5 years:<br>82 (51%) | Majority of participants<br>were age > 64 years:<br>6 (3.7%) | Majority of participants could be described as<br>vulnerable (e.g., HIV+, age under 6 months, care home<br>residents or transplant recipients): 82 (51%) |  |

NB: some countries were represented in multiple studies hence the country totals are > 162 and percentages > 100%. The counts of persons who were investigated were unclear in some studies, so that row does not add to 162.

**Table 2.**
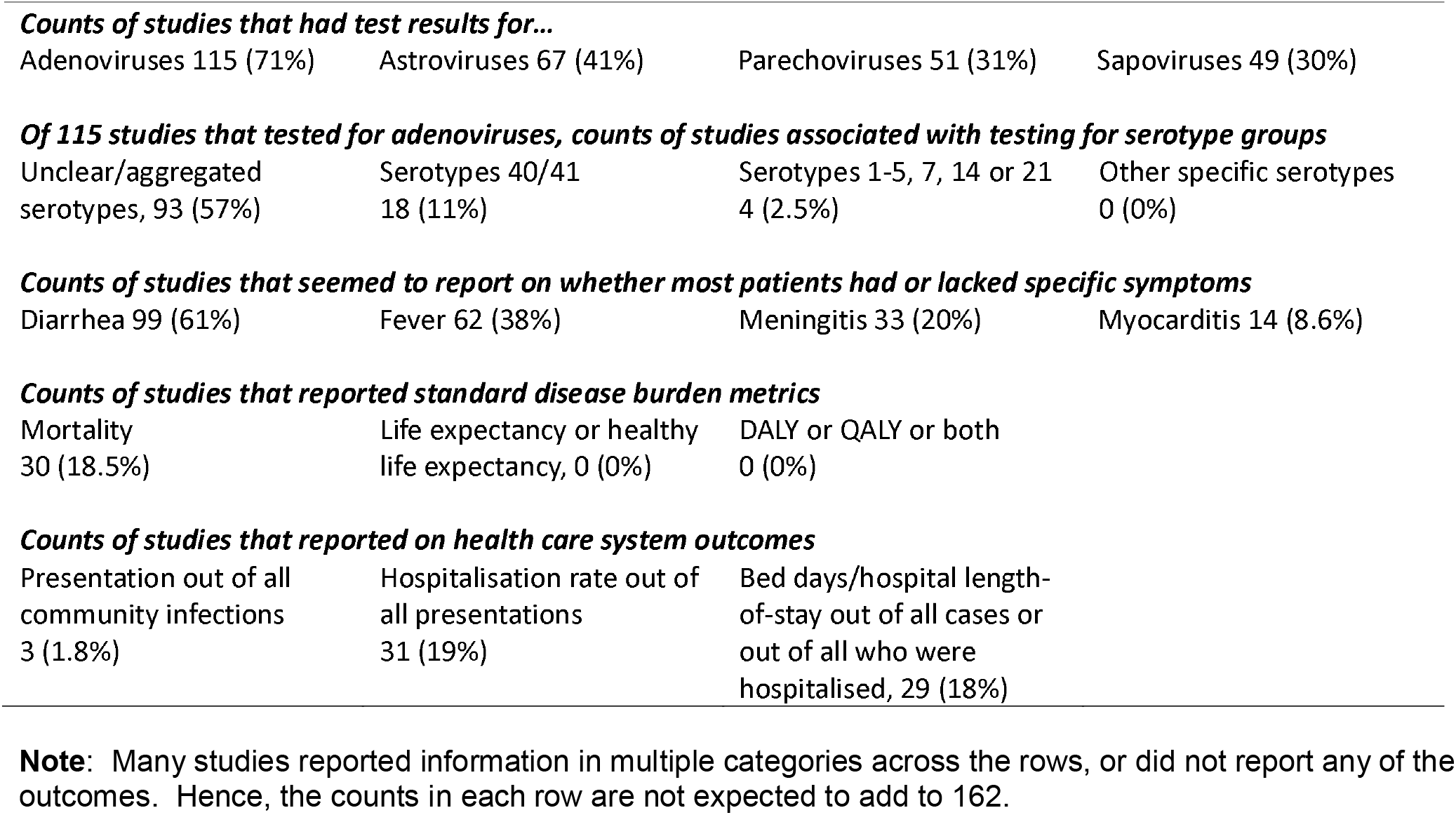
Summary of availability of information related to disease symptoms, disease burden or health care usage available in included studies.

|  |  |  |  |
| --- | --- | --- | --- |
| Adenoviruses 115 (71%) | Astroviruses 67 (41%) | Parechoviruses 51 (31%) | Sapoviruses 49 (30%) |
| ***Of 115 studies that tested for adenoviruses, counts of studies associated with testing for serotype groups*** |  |  |  |
| Unclear/aggregated serotypes, 93 (57%) | Serotypes 40/41 18 (11%) | Serotypes 1-5, 7, 14 or 21 4 (2.5%) | Other specific serotypes 0 (0%) |
| ***Counts of studies that seemed to report on whether most patients had or lacked specific symptoms*** |  |  |  |
| Diarrhea 99 (61%) | Fever 62 (38%) | Meningitis 33 (20%) | Myocarditis 14 (8.6%) |

***Counts of studies that reported standard disease burden metrics***
|  |  |  |
| --- | --- | --- |
| Mortality 30 (18.5%) | Life expectancy or healthy life expectancy, 0 (0%) | DALY or QALY or both 0 (0%) |

***Counts of studies that reported on health care system outcomes***
|  |  |  |
| --- | --- | --- |
| Presentation out of all community infections 3 (1.8%) | Hospitalisation rate out of all presentations 31 (19%) | Bed days/hospital length-of-stay out of all cases or out of all who were hospitalised, 29 (18%) |

**Note:** Many studies reported information in multiple categories across the rows, or did not report any of the outcomes. Hence, the counts in each row are not expected to add to 162.

**Figure 1.**
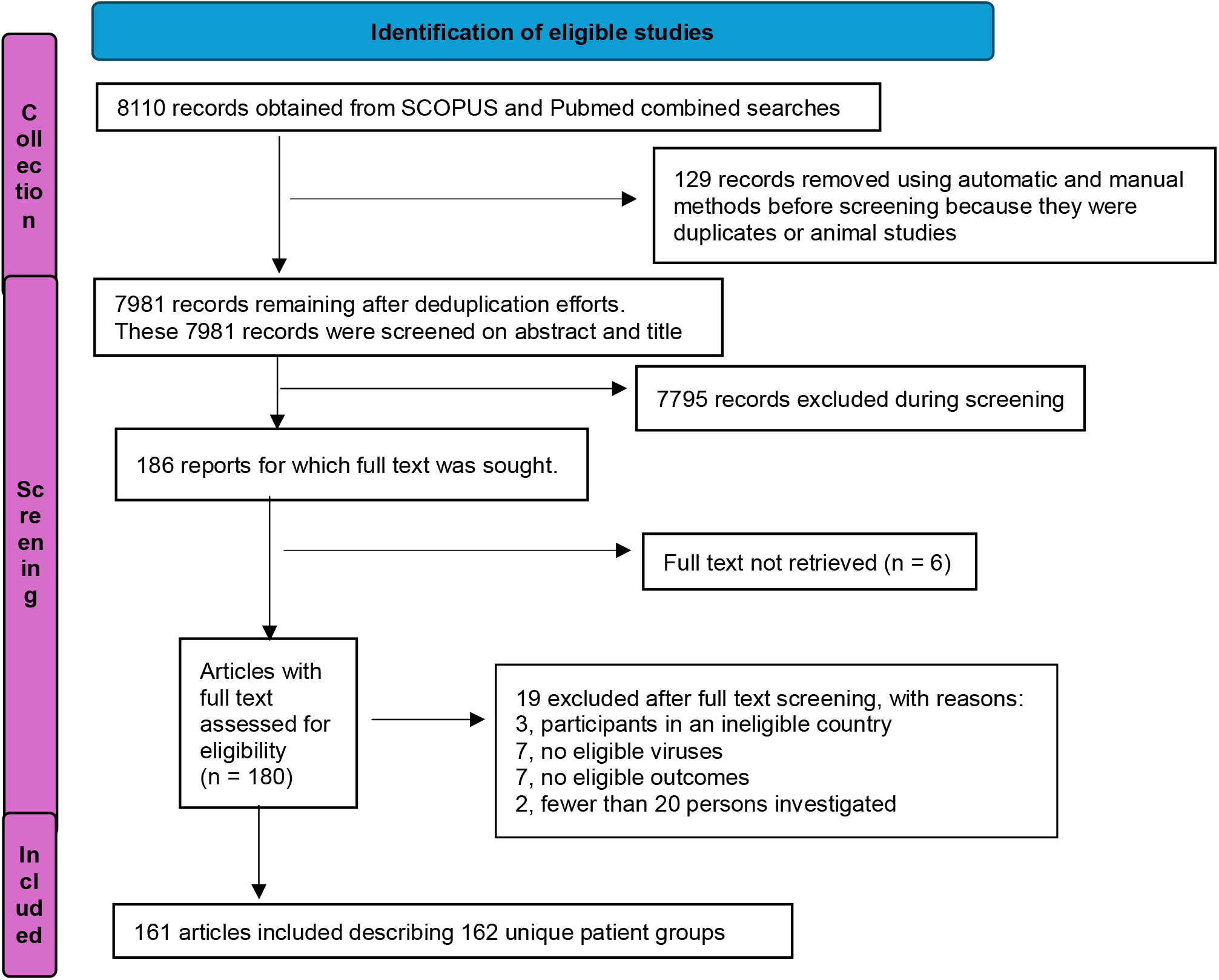
Study selection procedure

Table 1 shows that a small majority of studies were prospectively planned (n=91, 56%) and nearly all the studies were observational study designs (n=157, 97%). Prospective experimental studies were the most unusual study design (n=2, 1.2%). Germany was the single most represented country for participant location (n=37, 23%). About half (n=82, 51%) of the included studies were about viral presence in a population where the majority of patients were aged under 5 years old. About half of the studies (82, 51%) were focused on patients who could be classed as vulnerable. Typically, vulnerable status was assigned due to factors related to age (very young including recent premature birth or status as resident in care home for older persons), history as organ or tissue transplant recipient, HIV positivity, underlying comorbidities or congenital condition(s). Some patient populations had multiple reasons that caused them to be described as vulnerable. The median number of persons investigated for possible infection or infection outcomes was about 250.

Table 2 shows that tests for adenoviruses were much more frequent (in 71% of included studies) than tests for sapoviruses (30%), astroviruses (41%) or parechoviruses (31%). However, most studies (n=93, 57% of 115 studies that tested for adenoviruses) were not clearly specific about which enteric adenovirus spp./adenovirus serotype group was tested for (such as Serotype 40/41).

Table 2 also contains frequency counts for potential indicators of disease or health care system burden, such as morbidity measures and hospitalisation. Life expectancy calculations and the disease burden metrics DALYs and QALYs were not found in any included studies. Having diarrhoea or not was the only prespecified symptom information available in a majority of the 162 included studies. Health care system metrics such as hospitalisation rates or total bed days associated with infection status were available in at least 25 studies each.

NB: some countries were represented in multiple studies hence the country totals are > 162 and percentages > 100%. The counts of persons who were investigated were unclear in some studies, so that row does not add to 162.

## Discussion

Disease burden associated with infection by the target viruses has likely never been quantified and reported in scientific literature using the most validated disease burden metrics (QALYs or DALYs), and only rarely reported with an especially definitive complementary indicator of disease burden which is mortality rates. However, quantification of other specific morbidity indicators (eg., diarrhoea) or health care usage indicators (such as hospitalisation rates) that may be associated with infection status are feasible to synthesise from data in at least 29 original studies, even if the evidence were restricted only to the northwest European countries that we included.

We applied minimally biased and replicable methods that were transparently reported to quickly collect and summarise available literature that might support investigations of the disease burden associated with the target viruses. Because this was a rapid review, we did not achieve a fully comprehensive review, nor is a mapping review suitable for generating statements about actual and specific disease burdens associated with the target viral infections. Nevertheless, our study is novel. We are not aware of any previous structured systematic or comprehensive within a defined geographical area review that tried to map the evidence about disease burden associated with these target viruses. The information we report is valuable with respect to identifying the most documented disease and health service outcomes that can be associated with infection by the target viruses. This information can be used to design evidence syntheses that can inform future surveillance strategies with respect to gastrointestinal infections caused by the target virus genera.

The large body of primary studies we included, and the large counts of studies in countries included within and excluded from our review, suggests that there is widespread concern that the target viruses are often causing acute symptomatic gastrointestinal illness, important sequelae and generating frequent demand on health care systems. However, it is not clear yet that the target viruses are objectively “important” pathogens, where importance refers to either relative frequency as trigger for symptomatic illness or severity of definitively associated symptoms. The third study of Intestinal Disease in the UK (IID3; www.food.gov.uk/research/foodborne-pathogens/the-third-study-of-infectious-intestinal-disease-in-the-uk-iid3) reported that among 4062 stool samples collected between 2 Jan 2023 and 1 May 2025 from persons with symptomatic acute gastrointestinal illness, the majority (2627/4062) did not have a detected pathogen among 22 tested for [21]. Among our target viruses, sapovirus was found in 1.7% (n=71), astrovirus in 1.2% (n=50), and enteric adenoviruses in 0.4% (n=17) of the 4062 IID3 samples processed in this same 2023-2025 period. The target viruses were therefore linked with a relatively small proportion (3.3%) of acute gastrointestinal infections in this sample of UK persons who presented for health care because of symptoms suggestive of acute GI infection. The IID3 data do not indicate relative severity of symptoms nor prevalence of infection causing milder symptoms in the community. These observations are reminders that the availability of information in literature or databases about any specific potential gastrointestinal pathogen may not relate to actual relative importance as a cause of GI infection or morbidity in the wider population.

It is useful that our mapping review has established that much of the primary research into the target viruses describes fairly large cohorts. Large sample sizes can help make conclusions about specific outcomes more robust. It is also useful that we have documented there were at least 29 studies in populations that are highly relevant to the UK (north west Europe) that related infection status with frequency of specific symptoms (such as diarrhoea) or health care usage (e.g., hospitalisation rates among patients who presented for health care). With regard to investigating any apparent relationship between positivity and these symptoms or health service usage outcomes, we suggest that at least 20 primary research studies may be considered a minimum ‘good’ target count for generating a robust evidence summary. A study group size of at least 20 has greater potential to yield generalisable results, enable subgroup analysis, contain at least some studies that have low risk of bias and diversity in effect sizes, than a smaller study count. A primary study count higher than 20 may be desired depending on the synthesis methods that are applied to extracted data in systematic reviews. To achieve significance with a power of 0.80, it was estimated that approximately 27 studies are needed if using fixed effects meta-analysis and about 55 studies when using random effects meta-analysis [22]. However, we also note that many of the symptoms or health care usage outcomes that were less frequent in our rapid review (we found < 20 studies reporting these data), may be much more available to future reviewers if a wider geographic inclusion criteria (the rest of the world) was used. A decision to include studies from all countries in the world would need to consider comparability of diverse global populations, with respect to lifestyle, dietary and age-based demographic risk factors as well as variations in ascertainment. Variations in ascertainment are high over time as well as between countries. Our data collection exercise showed that distinctions in severity of illness harms should be possible between different adenovirus serotypes.

A mapping review helps to focus future research efforts on the most available evidence to explore with regard to a specific topic, such as understanding infectious GI disease burden caused by the target viruses. Any research question using systematic review methods to look at specific disease outcomes or health burden usage related to the target viruses will be better informed using the evidence mapping we have produced.

## Conclusion

DALYs or QALYs related to infectious status with sapoviruses, enteric adenoviruses, astroviruses or parechovirses has not been published with regard to populations in north west Europe. Estimates of measures of disease burden associated with these viral genera would be strengthened by community sampling to determine the asymptomatic colonisation rates of these same viruses in humans. Linking positivity status with frequency of some specific symptoms, health impacts or health care usage metrics is feasible using the existing evidence base. The most available outcome information is, among all persons who present for health care, hospitalisation rates and lengths of stay in hospital. It is possible to stratify by both virus genus and population location, including with respect to single large countries in north west Europe such as Germany or the UK.

## Key Points

- Modern test diagnostics mean that symptomatic illness is often linked with pathogens that have unknown total disease burden on human populations
- It is feasible to synthesize evidence about diarrheal symptoms or some health care usage metrics to infection with sapoviruses, astroviruses, parechoviruses or enteric adenoviruses
- There is little or no evidence linking disability-adjusted or quality-of-life-adjusted life-years to infection history with the target viruses
- Availability of information about specific severe symptoms or sequelae such as myocarditis or meningitis is also relatively limited for the target viruses

## Data Availability

Extracted data are available from lead author.

